# Analytical evaluation of thirty-two SARS-CoV-2 lateral flow antigen tests demonstrates sensitivity remains with the SARS-CoV-2 Gamma lineage

**DOI:** 10.1101/2021.12.08.21267455

**Authors:** Konstantina Kontogianni, Daisy Bengey, Dominic Wooding, Kate Buist, Caitlin Greenland-Bews, Christopher T. Williams, Margaretha de Vos, Camille Escadafal, Emily R. Adams, Thomas Edwards, Ana I. Cubas-Atienzar

## Abstract

The limit of detection (LOD) of thirty-two antigen lateral flow tests (Ag-RDT) were evaluated with the SARS-CoV-2 Gamma variant. Twenty-eight of thirty-two Ag-RDTs exceeded the World Health Organization criteria of an LOD of 1.0×10^6^ genome copy numbers/ml and performance was equivalent as with the 2020 B.1 lineage and Alpha variant.

---

The emergence of the variant of concern (VoC) Gamma (Pango P.1) began in November 2020 in the state of Amazonas in Brazil. The Gamma variant is estimated to be 1.7-2.4 times more transmissible than other local strains in Brazil (1) and quickly started to be detected at increasing rates from January 2021 onwards throughout the country, and became the predominant lineage associated with the second wave of infections with over 13 million confirmed cases and 350000 deaths (2). As of 30^th^ of November 2021, this strain has spread to 90 countries, and remains one of the most prevalent circulating in the Americas, with highest frequency in South America. In South America, the proportion of Gamma associated cases range from 5% to 100% depending on the country. Countries with higher levels of circulation of Gamma variant include Saint Vincent and Grenadines (100%), Haiti (100%), Trinidad and Tobago (50%), Argentina (30%) and Venezuela (30%) followed by Chile, Brazil, Peru, Ecuador, Suriname with 10%-20% of the circulating SARS-CoV-2 variants being Gamma (3).

The use of antigen SARS-CoV-2 lateral flow diagnostic tests (Ag-RDT) has become one of the first lines of defense against COVID-19, enabling early identification and isolation of cases to slow transmission, provision of targeted care, and protection of health system.

Gamma has 21 mutations, including 10 in the Spike (S) and 3 in the Nucleocapsid (N) proteins. As SARS-CoV-2 Ag-RDTs target S or N proteins, there is concern that these mutations could affect Ag-RDT performance. Thus, we sought to evaluate the LOD of 32 commercially available Ag-RDTs using the Gamma VoC, and to compare the results with previously determined LODs with Alpha and the ancestral B1 strain (5,7).

A clinical isolate from the P1 lineage of SARS-CoV-2 (hCoV-19/Japan/TY7-503/2021) was used for the evaluation. Briefly, the virus stock was propagated into Vero E6 cells (C1008; African green monkey kidney cells) that were maintained at 37°C with 5% CO_2_ in Dulbecco’s minimal essential medium (DMEM) supplemented with 4.5g/L glucose and L-Glutamine (Lonza, US), 10% foetal bovine serum (Sigma, US), and 50 units per ml of penicillin/streptomycin (Gibco, US). Frozen aliquots of the third passage of the virus were quantified via plaque assay as previously described (8).

The third passage was serially diluted in DMEM from 1.0×10^5^ to 1.0×10^2^ plaque forming units (pfu)/mL for determining the LODs of the 32 Ag-RDTs. The viral dilutions were added directly to the respective Ag-RDT extraction buffers, and Ag-RDTs were performed following the manufacturer instructions. Each dilution was tested in triplicate, with DMEM as a negative control. When the ten-fold LOD was found, two-fold dilutions were made and tested to confirm the lowest LOD (LLOD). The LOD was defined as the last dilution of a valid test where all three replicates were positive. Validity was determined by the presence of a control line, and only valid tests were included in the analysis. A positive result was interpreted visually by two operators by the presence of a test line. In the event of discordant result, a third operator read the test and acted as a tiebreaker.

RNA was extracted from each dilution using a QIAMP Viral RNA mini kit (Qiagen, Germany) and then genome copies (gcn)/mL were quantified using the COVID-19 Genesig RT-qPCR kit (PrimerDesign, UK) as previously described. We found 21/32 Ag-RDTs had an analytical LOD of ≤ 5.0 × 10^2^ pfu/ml (ActiveXpress, Bioperfectus, Core Test, Espline, Genedia, Fortress, iChroma, InTec, Joysbio, LumiraDx, Nadal, NowCheck, Panbio, PerkinElmer, RightSign, Roche, Standard F, Standard Q, Strong Step, Sure-Status and Wantai) fulfilling the British Department of Health and Social Care (DHSC) acceptable criteria. Additionally, 28/32 (including Biocredit, Covid-go, Excalibur, Mologic, Tigsun and Wondfo) had an LOD of ≤ 1.0 × 10^6^ gcn/ml fulfilling the recommendations in the WHO Target Product Profile for SARS-CoV-2 Ag-RDT (Table 1). The least sensitive tests with the Gamma variant were Innova, Flowflex, Hotgen, Onsite and RespiStrip.

**Table 1.**
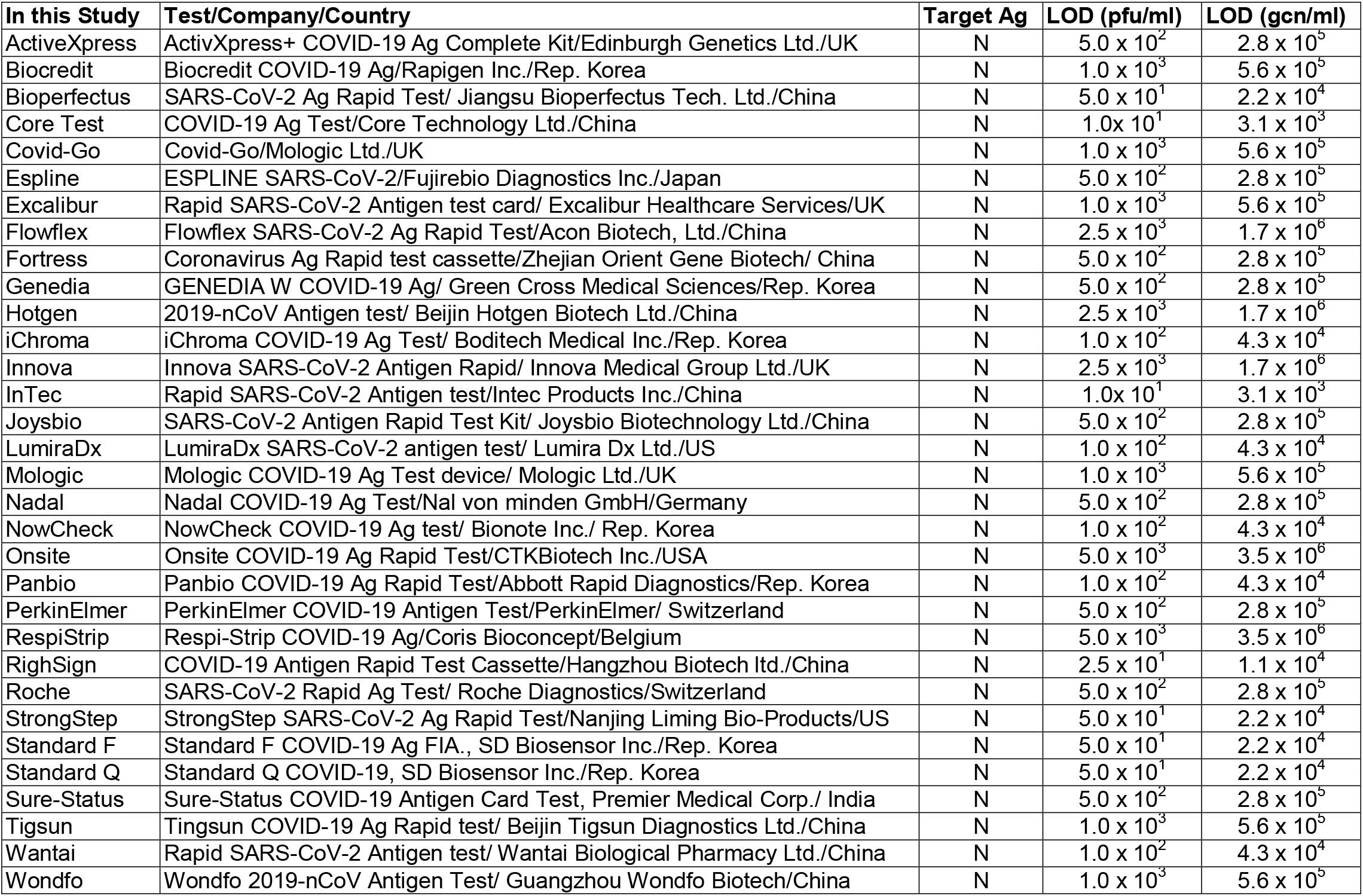
Description of the Ag-RDTs evaluated in this study and their LOD using the Gamma VoC.

Sensitivity to other SARS-CoV-2 strains was compared based on our previous work. There was no significant difference between the Ag-RDT LOD with Gamma in comparison to either the Alpha (Kruskal Wallis p = 0.315) or ancestral B.1 lineage (Kruskal Wallis p = 0.378) (Fig.1).

**Figure 1:**
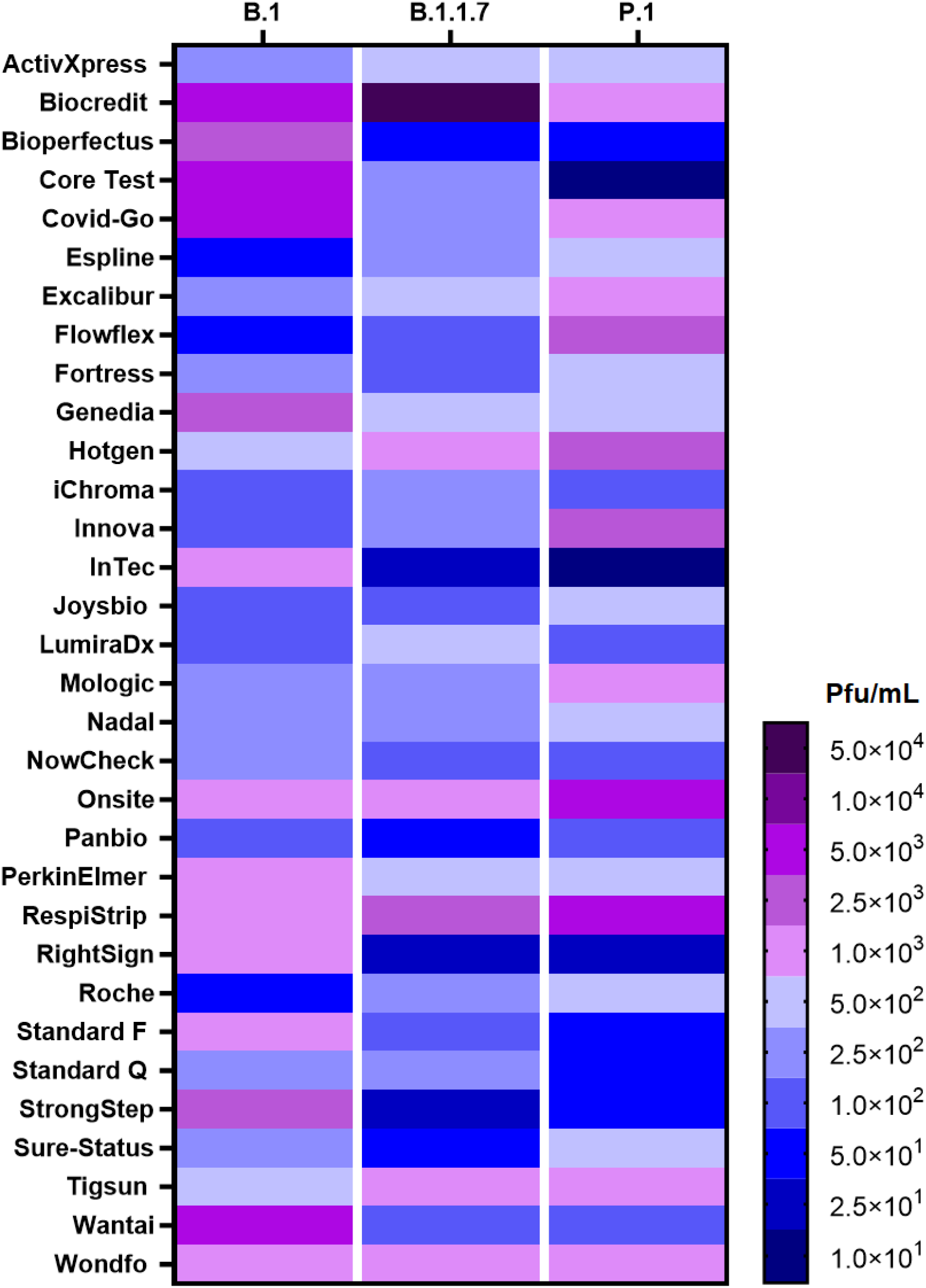
Heatmap comparing the LODs of 32 Ag-RDT using the Gamma (P.1), ancestral (B.1) and Alpha (B.1.1.7) variants. Data of the ancestral B.1 and Alpha partially taken from our previously published work (5,7). Blue colours indicated LODs fulfilling the DHSC and WHO criteria.

This is the most comprehensive analytical evaluation of COVID-19 Ag-RDTs with the Gamma VoC, and we have demonstrated that test performance is maintained, indicating Ag-RDT compatibility with this VoC. This evidence supports their continued usage in countries where the Gamma strain is circulating. However, clinical diagnostic evaluations in prospective cohorts in these localities are required to provide definitive data on their clinical performance. All the Ag-RDTs evaluated in this study detect the N protein, which contains fewer mutations than S in the Gamma lineage. Ag-RDTs targeting S may be affected differently. It is vital that diagnostic evaluations continue to monitor test performance in emerging variants, to ensure continued diagnostic performance.

## Data Availability

Data is available upon request

## Acknowledgements

We thank Grant L Hughes for facilitating the SARS-CoV-2 isolate.

## About the author

Konstantina Kontogianni is a research technician at the Liverpool School of Tropical Medicine, and her primary interest is the diagnosis of tuberculosis and respiratory viruses including SARS-CoV-2.

